# Predicting daily sleep outcomes from continuous HRV in female chronic pelvic pain disorders

**DOI:** 10.64898/2026.07.16.26357390

**Authors:** Rhyann Clarke, Samia Shahnawaz, Robert Hirten, Jovita Rodrigues, Kyle Landell, Matteo Danieletto, Gerard Ona, Ipek Ensari

## Abstract

**Background:** Female chronic pelvic pain disorders (CPPDs) are highly prevalent and frequently accompanied by sleep disturbance and autonomic nervous system (ANS) dysregulation. Heart rate variability (HRV), a non-invasive index of ANS function, may provide an objective, physiological correlate of sleep health and can be monitored using wearable devices, enabling a continuous, scalable approach.

**Objectives:** This study examined whether wearable-derived daily HRV metrics are associated with self-reported sleep disturbance in women with CPPD(s) compared with healthy controls, using epoch-level data and generalized additive models.

**Methods:** We conducted a retrospective observational study using up to 90 days of data from a mobile health research app. Participants were 128 women with CPPD(s) and 63 demographically matched healthy controls, who completed a daily PROMIS-based 3-item sleep disturbance questionnaire and wore Fitbit devices that provided 5-minute HRV epochs. Primary predictors were high frequency (HF) and low frequency (LF) power and root mean square of successive differences (RMSSD), with group (CPPD vs control), daily pain severity, and menstrual status as covariates. We fit separate generalized additive mixed models (GAMMs) for each HRV metric with a nonlinear smooth term and an HRV × Group interaction.

**Results:** Higher HF and RMSSD were associated with lower sleep disturbance scores, and these associations were stronger in controls than in the CPPD group (HF × group B ≈ –1.59, p < 0.00010; RMSSD × group B ≈ –0.58, p < 0.0001). LF showed a more complex pattern but also differed by group (B ≈ -0.531, p < 0.0001). HRV smooth terms were highly nonlinear, and models explained ~8–9% of deviance in sleep disturbances. Pain severity and menstrual bleeding were strongly associated with worse sleep.

**Conclusion:** These findings indicate small but consistent associations between wearable-derived HRV metrics and daily sleep disturbances in women with CPPD(s) and healthy controls, with weaker associations in CPPD(s). Integrating continuous HRV with symptom tracking could support low-burden and multimodal monitoring of sleep health in chronic pelvic pain, but prospective validation is needed before HRV can be used for diagnostic or treatment response decision making.

## Introduction

Women with chronic pelvic pain disorders (CPPD(s); e.g., endometriosis, adenomyosis, fibroids, bladder pain syndrome) experience persistent or recurrent pain in the pelvis or lower abdomen that interferes with daily functioning and often remains inadequately managed over years. CPPD(s) affect an estimated 5–26% of women worldwide and are associated with reduced quality of life, impaired productivity, and high health care utilization ^1^. Despite this burden, many patients face diagnostic delays of 7-10 years and limited relief from available treatments ^1^. Sleep disturbance is one of the most common and disabling comorbidities in CPPD(s), with a reported prevalence of 72% and elevated rates of co-morbid depression and anxiety ^2^. Because sleep plays a central role in pain processing, emotional regulation, and overall health, its disruption may further exacerbate both pain and psychosocial burden in CPPD(s) ^3^.

In women with chronic pelvic pain, poor sleep has been linked to higher pain intensity, worse emotional functioning, and increased interference with daily activities ^4^. Yet sleep symptoms are often neglected in clinical encounters that prioritize acute pain and reproductive outcomes ^2^. Existing evidence also suggests that CPPD patients carry a disproportionate burden of mood and anxiety symptoms, and that sleep disturbance is intertwined with these mental health outcomes ^2^. Characterizing how sleep disturbances fluctuate in relation to physiological state in CPPD(s) is therefore a critical step toward more comprehensive, patient-centered care. A growing body of work implicates autonomic nervous system (ANS) dysregulation as one plausible shared mechanism linking chronic pain and sleep disruption ^1,2^ . Women with CPPD(s) demonstrate altered autonomic function, including reduced vagal activity and a shift toward sympathetic dominance, compared with healthy controls ^5^. Heart rate variability (HRV), a non-invasive index of beat-to-beat variation in heart period, provides a window into ANS regulation of cardiovascular and stress responses. HRV has time- and frequency domains that have been indicated to be relevant for health outcomes.

In general, HRV time-domain measurements decline with decreased health, and a person’s baseline is influenced by their age, mental health, and physical disorders ^6^. In healthy cohorts, lower HRV during sleep, particularly reduced high frequency (HF) power and time-domain metrics such as the root mean square of successive differences (RMSSD), has been prospectively associated with more sleep complaints, worse stress regulation, and increased risk of cardiometabolic diseases ^7^,^8^. RMSSD quantifies fluctuations in time intervals between consecutive heartbeats, which can indicate how “deep” a person’s sleep is ^9^. RMSSD is primarily associated with the parasympathetic nervous system’s influence on heart rate and reflects the body’s ability to recover from various stressors. HF also represents parasympathetic activity during sleep, and higher HF often reflects better sleep quality ^6^. Low frequency (LF) reflects mixed sympathetic and parasympathetic activity and is therefore more difficult to interpret, but lower LF has been seen to be associated with higher sleep scores ^10^. However, most of this evidence comes from general or cardiometabolic populations, and little is known about how continuous HRV relates to sleep disturbances specifically in women with CPPD(s) ^11^.

In routine care, sleep is typically assessed using brief questionnaires or single item ratings obtained during clinical visits. Although these instruments are practical, they are vulnerable to recall bias, cannot capture night to night variability, and rarely capture the temporal coupling between sleep, pain, and physiology [5]. Wearable devices enable continuous, low-burden collection of HRV and related physiological signals in real-world settings. Evidence from validation studies demonstrates that consumer grade wearables such as the Fitbit trackers produce nocturnal HRV metrics with acceptable agreement to reference devices ^12^. Integrating these data streams with daily self-reports could help delineate how ANS dynamics relate to sleep disturbances in CPPD(s) ^13^. Mobile health (mHealth) Apps offer an opportunity to pair daily patient reported outcomes with continuous wearable data to characterize symptom-physiology relationships in CPPD(s) at scale. Such information could inform more nuanced digital monitoring strategies and hypothesis-driven intervention designs for this underserved population.

Accordingly, the primary objective of this study was to examine the association between HRV metrics (HF, LF, and RMSSD) and daily sleep disturbance among those with CPPD(s), and to test whether these associations differ between those with versus without CPPD(s). Using 90 days of user-tracked sleep and wearable-tracked HRV data, we investigated (1) whether higher parasympathetic indexed HRV metrics would be associated with fewer sleep disturbances; (2) whether the magnitude of these associations would differ in CPPD compared with controls; and (3) how much additional variance in daily sleep disturbance is captured by HRV beyond pain and menstrual status.

## Methods

### Study Design & Procedures

This study was a retrospective analysis of data collected over 90 days through an ongoing observational mHealth study of adults with a female chronic pelvic pain disorder (CPPD(s); e.g., endometriosis, adenomyosis, fibroids). The parent study was designed to capture patient generated health data with high temporal complexity using a research app (ehive) and wearable trackers under ecologically valid conditions ^14^. Participants completed app-based symptom tracking and wore Fitbit devices continuously during the study period. Fitbit-derived sleep windows were used to identify sleep periods based on proprietary estimates from movement and heart rate patterns, and HRV data were extracted as repeated 5-minute epochs. This secondary analysis focused on the association between epoch-level HRV and daily self-reported sleep disturbances. The study protocol was approved by the Institutional Review Board of the Icahn School of Medicine at Mount Sinai, and all participants provided informed consent before enrollment (IRB # STUDY-23-00721). Study procedures were conducted in accordance with the Declaration of Helsinki. Participants were onboarded remotely to the study app and device procedures and were instructed to complete symptom tracking and wear the Fitbit device throughout the 90-day study period.

### Study Participants

The analytic sample included 128 women with CPPD(s) and 63 demographically matched controls who all contributed app-based sleep data and wearable HRV data during the study period. The participant flow can be seen in Supplemental Figure 1.

Participants in the CPPD(s) group were enrolled based on self-reported clinician diagnosed chronic pelvic pain disorder(s), while controls were recruited to provide a comparison group without a CPPD. Eligibility was restricted to individuals assigned female at birth with female reproductive anatomy who menstruate, were between 18 and 64 years of age, and were able to read and write in English. Exclusion criteria included current pregnancy, recent childbirth, planned pregnancy during the study period, and major active comorbid medical conditions that could interfere with participation or autonomic measurements. Participants were recruited from Mount Sinai Health System and affiliated clinical or community channels and were enrolled between April 2023 and July 2025.

### Data sources

Participants used the ehive App to report daily sleep outcomes and related symptoms and wore Fitbit devices to passively collect physiological data. HRV data were available as repeated 5-minute epochs, yielding an epoch-level dataset nested within person-days and participants. Each HRV epoch was linked to the same day’s self-reported sleep disturbance score, such that multiple HRV observations contributed to a single daily outcome per participant. The final analytic dataset included 410,180 epoch-level observations across all participants and study days. Analyses were restricted to days with concurrent HRV and self-tracked sleep data.

### Sleep outcome

The primary outcome was a daily composite sleep disturbance score derived from 3 day-level ehive app-based questions (See Supplemental File). The 3-item daily sleep disturbance composite indicated good internal consistency in the analytic sample (Cronbach’s alpha = 0.80, standardized alpha = 0.80; average inter-item correlation = 0.57). Higher scores on each item (reverse-coded where necessary) reflected greater sleep disturbance, and all items demonstrated strong corrected item–total correlations (r = 0.54–0.76), supporting retention in a single summed 0–12 scale. In mixed-effects and generalized additive models, the daily composite sleep scores was strongly and positively associated with PROMIS Sleep Disturbance raw and T scores (all p < 0.001; See Supplementary Tables 1-3 for complete results), accounting for approximately 39% of the variance in PROMIS sleep outcomes, which supports convergent validity of the daily measure.

### HRV predictors

The primary predictors were three HRV metrics obtained from Fitbit data: HF power, LF power, and RMSSD. HF and RMSSD were selected as indices with stronger parasympathetic interpretation, whereas LF was included as a broader index reflecting mixed autonomic influences. HRV was analyzed at the 5-minute epoch level to preserve temporal variation in autonomic physiology rather than reducing each day to a single summary value. We fit separate models for HF, LF, and RMSSD to avoid multicollinearity and allow interpretation of metric specific associations with the sleep outcome.

### Covariates

The primary covariates were daily pain severity and menstrual cycle status. Pain severity was included to account for the well-established coupling between pain and sleep disturbances in CPPD, while menstrual status was included as a time-varying physiological covariate that could influence both sleep and autonomic function.

### Statistical design and analysis

Our primary analyses focused on concurrent associations between nocturnal HRV and same-night self-reported sleep disturbance. This choice reflects the core aims of the study: (1) to evaluate whether wearable-derived HRV can serve as a passive proxy for daily sleep disturbance in circumstances where self-tracking is unavailable or incomplete, and (2) to establish the psychometric coherence and construct validity of a brief daily sleep disturbance composite. Concurrent HRV–sleep coupling is most relevant to estimating missing or low-burden daily. All analyses were conducted using generalized additive mixed models (GAMMs) implemented in the mgcv package in R. ^15^. GAMMs were chosen because they flexibly accommodate nonlinear associations while accounting for the repeated-measures structure of the data ^16^. For each HRV metric, the daily sleep disturbance score was regressed on a smooth term for the standardized HRV metric, a linear HRV x group interaction, a smooth term for pain severity, menstrual status, and a participant-level random intercept. We used the smooth term for HRV to allow estimation of the potentially nonlinear associations between HRV level and sleep disturbance. The interaction term between Group status (CPPD vs control) and HRV was included to test whether HRV-sleep associations differed by clinical group. Participant and day number were included as random intercepts to account for repeated observations within individuals and the nested structure of the HRV epochs within days.

### Model specifications and diagnostics

Continuous HRV predictors were standardized prior to modeling to improve comparability across metrics and stabilize model fitting. The basis dimension k for the HRV smooth was set a priori for each model (HF: k=12; LF: k=10; RMSSD: k=10) to allow sufficient flexibility while limiting overfitting. Smoothing parameters were estimated using penalized likelihood as implemented in mgcv, and model adequacy was evaluated using standard GAM diagnostics, including inspection of effective degrees of freedom and evaluation of whether the chosen basis dimensions were adequate. The GAMMs estimate nonlinear associations between epoch-level HRV values and the daily sleep disturbance score, while accounting for repeated measures within participants via a random intercept. We used the adjusted R^2^, deviance explained, generalized cross validation (GCV), and scale estimates to evaluate model fit.

## Results

### Participant Demographics

The participant demographic characteristics are provided above in Table 1. The analytic sample included 128 women with CPPD(s) and 63 controls. The mean age was 37 years in the CPPD group and 35 years in the control group. There were no significant demographic differences between participants with versus without CPPD(s). Most participants were white, college educated, and employed, and most were recruited through Mount Sinai Health System (MSHS).

**Table 1.**
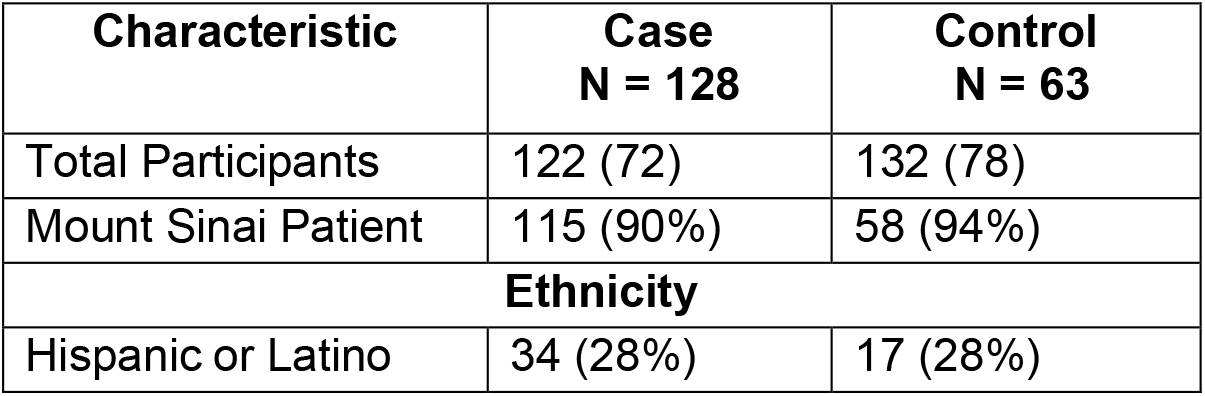
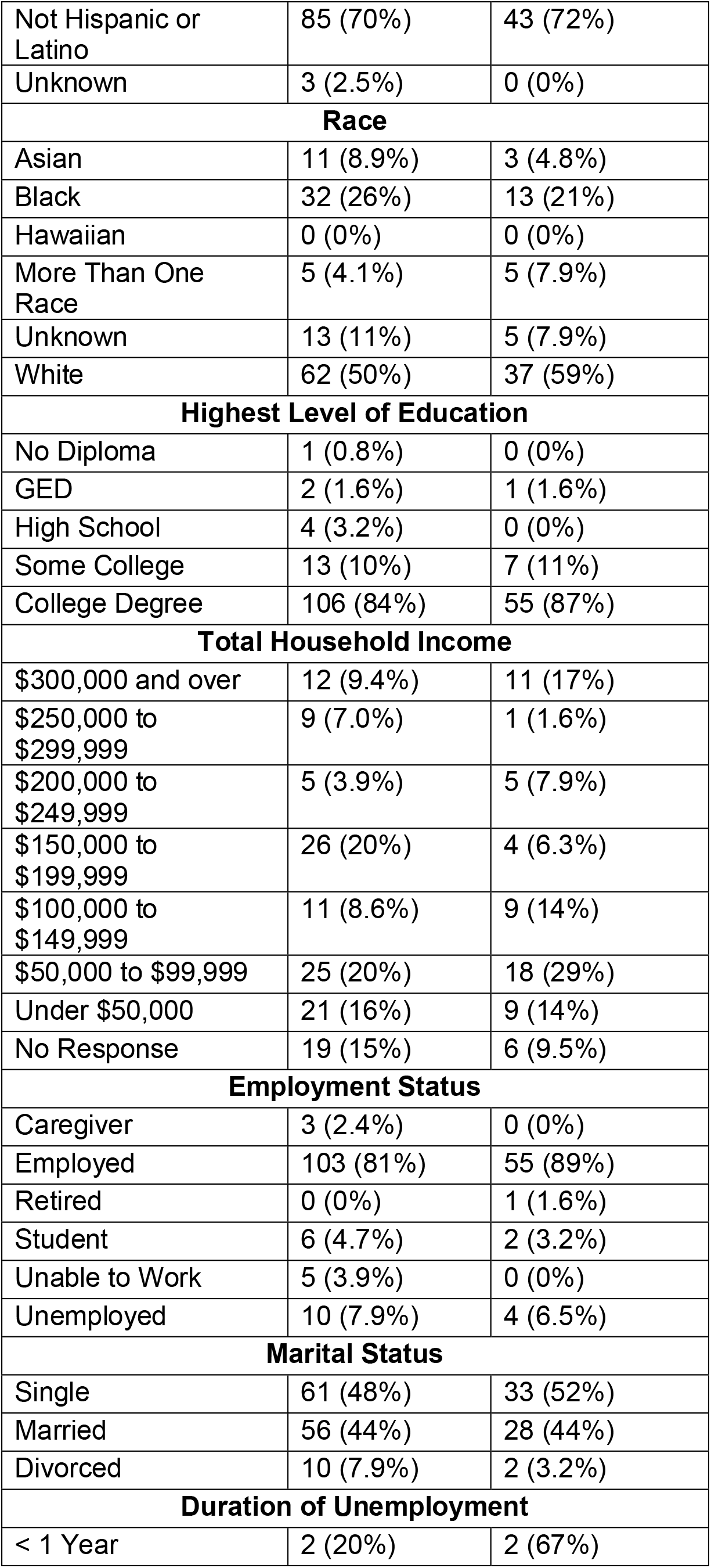
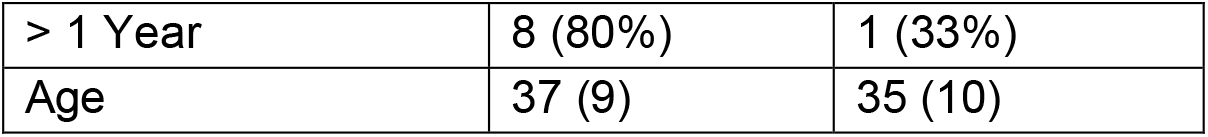
Participant Demographics.

**Table 2.** Parametric Effects from LF GAMM.

| Model | Predictor | Estimate | se | t | p |
| --- | --- | --- | --- | --- | --- |
| LF | (Intercept) | -0.541 | 0.014 | -37.421 | <0.0001 |
|  | LF Standardized | -0.124 | 0.014 | -8.932 | <0.0001 |
|  | Control group | -1.005 | 0.028 | -35.871 | <0.0001 |
|  | Pain severity | 0.320 | 0.002 | 142.278 | <0.0001 |
|  | Menstrual period (yes v. no) | -0.450 | 0.012 | -36.809 | <0.0001 |
|  | LF standardized x Control group | -0.531 | 0.041 | -12.830 | <0.0001 |

**Table 3.** Parametric Effects from HF GAMM.

| Model | Predictor | Estimate | se | t | p |
| --- | --- | --- | --- | --- | --- |
| HF | (Intercept) | 0.832 | 0.012 | 70.543 | <0.0001 |
|  | HF standardized | 0.129 | 0.022 | 5.874 | <0.001 |
|  | Control group | -1.471 | 0.036 | -40.502 | <0.0001 |
|  | Menstrual period (yes v. no) | -0.407 | 0.012 | -33.146 | <0.0001 |
|  | HF standardized x Control group | -1.600 | 0.197 | -8.138 | <0.001 |

**Table 4.** Parametric Effects from RMSSD GAMM.

| Model | Predictor | Estimate | se | t | p |
| --- | --- | --- | --- | --- | --- |
| RMSSD | (Intercept) | 0.836 | 0.012 | 69.963 | <0.0001 |
|  | RMSSD standardized | 0.150 | 0.069 | 2.190 | 0.0285 |
|  | Control group | -1.424 | 0.031 | -45.930 | <0.0001 |
|  | Menstrual period (yes v. no) | -0.390 | 0.012 | -31.702 | <0.0001 |
|  | RMSSD standardized x Control group | -0.570 | 0.069 | -8.269 | <0.0001 |

**Table 5.** Results of the GAMM smooth terms.

| Model | Smooth term | edf | ref_df | F | p |
| --- | --- | --- | --- | --- | --- |
| LF | Standardized LF | 8.040 | 8.433 | 32.050 | <0.0001 |
| LF | Participant ID | 1.000 | 1.000 | 4161.620 | <0.0001 |
| LF | Study day | 0.991 | 1.000 | 166.060 | <0.0001 |
| HF | Standardized HF | 10.004 | 10.419 | 79.090 | <0.0001 |
| HF | Pain severity | 8.887 | 8.995 | 2677.150 | <0.0001 |
| HF | Participant ID | 1.000 | 1.000 | 5476.810 | <0.0001 |
| HF | Study day | 0.995 | 1.000 | 209.690 | <0.0001 |
| RMSSD | Standardized RMSSD | 8.262 | 8.491 | 40.010 | <0.0001 |
| RMSSD | Pain severity | 8.896 | 8.996 | 2683.200 | <0.0001 |
| RMSSD | Participant ID | 1.000 | 1.000 | 5459.020 | <0.0001 |
| RMSSD | Study day | 0.995 | 1.000 | 219.620 | <0.0001 |

### Primary HRV Models

Across all three models, controls reported lower sleep disturbance than participants with a CPPD, and daily pain severity and menstrual status were consistently associated with sleep disturbance. In the LF model, controls had on average 1.01 points lower sleep disturbance scores than CPPD participants (B = −1.01, SE = 0.03, p < 0.001). A 1-unit increase in daily pain severity was associated with a 0.32-point increase in sleep disturbance (SE = 0.00, p < 0.001), and menstrual status was also associated with differences in sleep disturbance (B ≈ −0.45, SE = 0.01, p < 0.001). Similar group differences were observed for HF (B = −1.47, SE = 0.04, p < 0.001) and RMSSD (B = −1.42, SE = 0.03, p < 0.001), with menstrual status remaining a predictor in both models (HF: B ≈ −0.41, SE = 0.01; RMSSD: B ≈ −0.39, SE = 0.01; all p < 0.001).

LF showed a nonlinear association with sleep disturbance, and the linear LF-by-group indicated that the association differed by group. In CPPD participants, a 1 SD increase in LF was associated with a modest reduction in sleep disturbance (B = −0.12, SE = 0.01, p < 0.001), whereas in controls the corresponding linear effect was larger (B ≈ −0.66, combining the main LF and interaction terms; LF x group interaction B = −0.53, SE = 0.04, p < 0.001). The LF smooth term was highly nonlinear (EDF ≈ 8.0 of 10, F ≈ 32.1, p < 0.001), indicating a curved relationship between LF levels and sleep disturbance.

HF and RMSSD showed similar group differences, with stronger inverse associations in controls than in CPPD(s). In CPPD participants, a 1 SD increase in HF was associated with a small increase in sleep disturbance (B = 0.13, SE = 0.02, p < 0.001), while in controls it was associated with a substantial decrease (B ≈ −1.47; HF × group interaction B = −1.60, SE = 0.20, p < 0.001). The HF smooth term was strongly nonlinear (EDF ≈ 10.0 of 12, F ≈ 79.1, p < 0.001). For RMSSD, a 1 SD increase corresponded to a small increase in sleep disturbance in CPPD(s) (B = 0.15, SE = 0.07, p = 0.029) but a decrease in controls (B ≈ −0.42; RMSSD X group interaction B = −0.57, SE = 0.07, p < 0.001), with the RMSSD smooth again clearly nonlinear (EDF ≈ 8.3 of 10, F ≈ 40.0, p < 0.001).

Participant- and day level random effects were highly significant in all models (participant EDF ≈ 1.0, F range 4161.6–5476.8; day EDF ≈ 1.0, F range 166.1–219.6; all p < 0.001), indicating substantial heterogeneity in baseline sleep disturbance between individuals and across days.

## Discussion

In this study, we leverage a disease-specific, day-level sleep disturbance score and high-frequency HRV sampling using consumer wearables to characterize autonomic–sleep associations in women with CPPD(s) and matched controls under real-world conditions. By combining a daily PROMIS-based sleep measure with 5-minute HRV epochs and generalized additive mixed models, this work moves beyond one-off clinic questionnaires and shows how autonomic state relates to sleep disturbance in CPPD(s) across days. Unlike most CPPD studies that treat sleep as a secondary binary or single-item outcome, we captured continuous, within-person variability in sleep disturbance and related it to repeated physiological measures, while adjusting for pain and menstrual status. To our knowledge, this is among the first CPPD studies to (1) use a day-level, disease-specific sleep disturbance score as the primary outcome, (2) model nonlinear associations between multiple HRV metrics and sleep disturbance, and (3) explicitly account for both participant- and day-level random effects in a wearable-based dataset.

We note two consistent findings across the HF, LF, and RMSSD models. First, control participants had substantially lower sleep disturbance scores than CPPD participants (1–1.5 scale points at baseline). This highlights a clinically meaningful sleep burden associated with CPPD even after accounting for pain. This finding aligns with prior work linking chronic pelvic pain to poorer sleep and worse functioning compared with pain-free controls ^4^. It also stays consistent with broader chronic pain research documenting that sleep disturbances are both highly prevalent and strongly tied to disability and mood symptoms ^17^. Second, pain severity had a strong and nonlinear association with sleep disturbance, reinforcing the central role of pain in the sleep–pain dyad in CPPD. The nonlinearity indicates that participants are affected by pain differently, and this is supported with the large contribution with participant random effect. Longitudinal and experimental work in chronic pain suggests that sleep disturbance may be a stronger predictor of next-day pain than the reverse, and that treating sleep can improve pain outcomes ^18,19^. These results support treating sleep disturbance as a core target and outcome in CPPD care, rather than a secondary symptom.

Higher LF was associated with lower sleep disturbance, particularly in controls, with a modest effect in CPPD(s) and a steeper favorable slope in controls. Similarly, higher HF and higher RMSSD were associated with lower sleep disturbance among healthy control participants. For participants with CPPD(s), however; the linear component of the association was flat or even slightly adverse. Nonlinear smooths for all three metrics were statistically significant, with effective degrees of freedom near the upper bound, indicating curved relationships that change across the HRV range rather than simple monotone trends. These findings collectively suggest that higher parasympathetic indexed HRV is associated with fewer sleep disturbances in healthy participants. This is consistent with prior work linking higher

HF-HRV to fewer sleep complaints and better stress regulation in working adults ^7^. In contrast, women with CPPD(s) appear to have a blunted or altered coupling between HRV and perceived sleep disturbance. This may reflect chronic autonomic dysregulation that weakens the usual buffering role of vagal activity ^11^. These group differences are consistent with evidence that chronic pain states are characterized by reduced parasympathetic activity and relative sympathetic dominance, as seen in chronic musculoskeletal pain and dysmenorrhea ^20^ and with CPPD-specific work ^21^ showing autonomic symptom burden and altered HRV profiles relative to controls ^5^. Current findings demonstrating that, wherein CPPD(s), HRV is still significantly associated with sleep disturbance but in a way that is weaker and more complex than in controls. This may reflect a combination of persistent autonomic arousal, hormonal influences, and central sensitization. Together, these factors may decouple day-to-day vagal fluctuations from subjective sleep disturbance. This also suggests that HRV-based interventions or feedback, such as HRV biofeedback or breathing exercises, might benefit sleep in CPPD(s), although condition-specific tailoring may be needed ^18^.

The daily PROMIS-derived sleep disturbance score is another important contribution. Standard tools such as the PSQI, Insomnia Severity Index, or DSM-5 PROMIS Level-2 Sleep Disturbance measure are typically designed for longer recall windows and are not well suited for dense day-level sampling or digital phenotyping ^22,2324^. By adapting a brief sleep measure for daily use, we were able to capture within-person fluctuations in sleep disturbance over time and examine how these changes align with pain and wearable-derived physiology. In our sample, the daily composite demonstrated good internal consistency and concurrent validity based on comparison to PROMIS scores. Moreover, higher composite scores were associated with higher pain severity, menstrual bleeding days, and lower parasympathetic-indexed HRV in expected directions. This indicates that the daily sleep outcome behaves as predicted within a broader network of theoretically related constructs and thereby demonstrating satisfactory nomological validity ^25–27^. Taken together, this day-level compositive sleep measure provides a psychometrically coherent and clinically interpretable outcome for tracking subjective sleep disturbance in CPPD(s) ^13,28^.

This measurement approach also has implications for the design of digital interventions in CPPD. The robust and nonlinear associations of pain and menstrual status with sleep disturbance highlight the importance of capturing both symptom burden and hormonal context in mHealth tools ^22^. Coupling day-level sleep measures with passively collected HRV may help identify high-risk nights or flare ups, inform the timing of behavioral sleep interventions, or personalized recommendations around activity, relaxation, and analgesia. Given the evidence that improving sleep can attenuate pain and that higher HRV may reflect better pain-inhibition capacity ^11^, integrating HRV informed prompts (e.g., suggesting wind-down routines on days with low HRV and high pain) is a plausible direction for just-in-time interventions for CPPD(s) ^28,29^. However, prospective trials will be needed to test whether HRV-augmented digital interventions meaningfully improve sleep and pain outcomes beyond symptom-only tracking.

This study has several limitations. First, the observational design precludes causal inference about whether altered HRV and sleep disturbance are a consequence of CPPD, a maintaining factor, or both. The models quantify associations between epoch-level HRV and day-level sleep disturbance but cannot determine temporal ordering or directionality. Second, sleep disturbance was assessed via brief self-report rather than PSG; while daily PROMIS-based measures are well suited for capturing subjective disturbance, they do not index sleep architecture or specific disorders such as sleep-disordered breathing. Third, HRV metrics were derived from Fitbit devices. Although Fitbits show reasonable accuracy for time-domain measures like RMSSD, they may yield more variable frequency-domain estimates (HF and LF) and are susceptible to proprietary preprocessing and movement of artifacts. Finally, the sample was drawn from a single health system and self-selected into a digital study, which may limit generalizability to more diverse populations or to individuals with limited access to technology.

## Conclusion

We herein demonstrate that wearable-derived HRV metrics are nonlinearly and differentially associated with daily sleep disturbance in women with CPPD(s) and matched controls. Higher parasympathetic-indexed HRV (HF and RMSSD) and higher LF are robustly linked to lower sleep disturbance in controls, whereas these associations are attenuated and more complex in CPPD(s), with contributing effects of pain and menstrual status. Although HRV explains only a modest portion of variance in sleep disturbance, integrating day-level, disease-specific sleep measures with continuous HRV and symptom data offers a promising, low-burden strategy for characterizing sleep health and autonomic dysregulation in CPPD(s). Future work should build on these findings to develop and test multimodal digital tools that treat sleep disturbance as a core target in CPPD care and that leverage HRV as one component of personalized monitoring and intervention rather than as a stand-alone biomarker.

## Data Availability

The data generated in the present study can be made available from the corresponding author after the parent grant period is over upon reasonable request, provided that the request is consistent with the terms of the informed consent.

## Supplemental Files

**Supplemental Figure 1.**
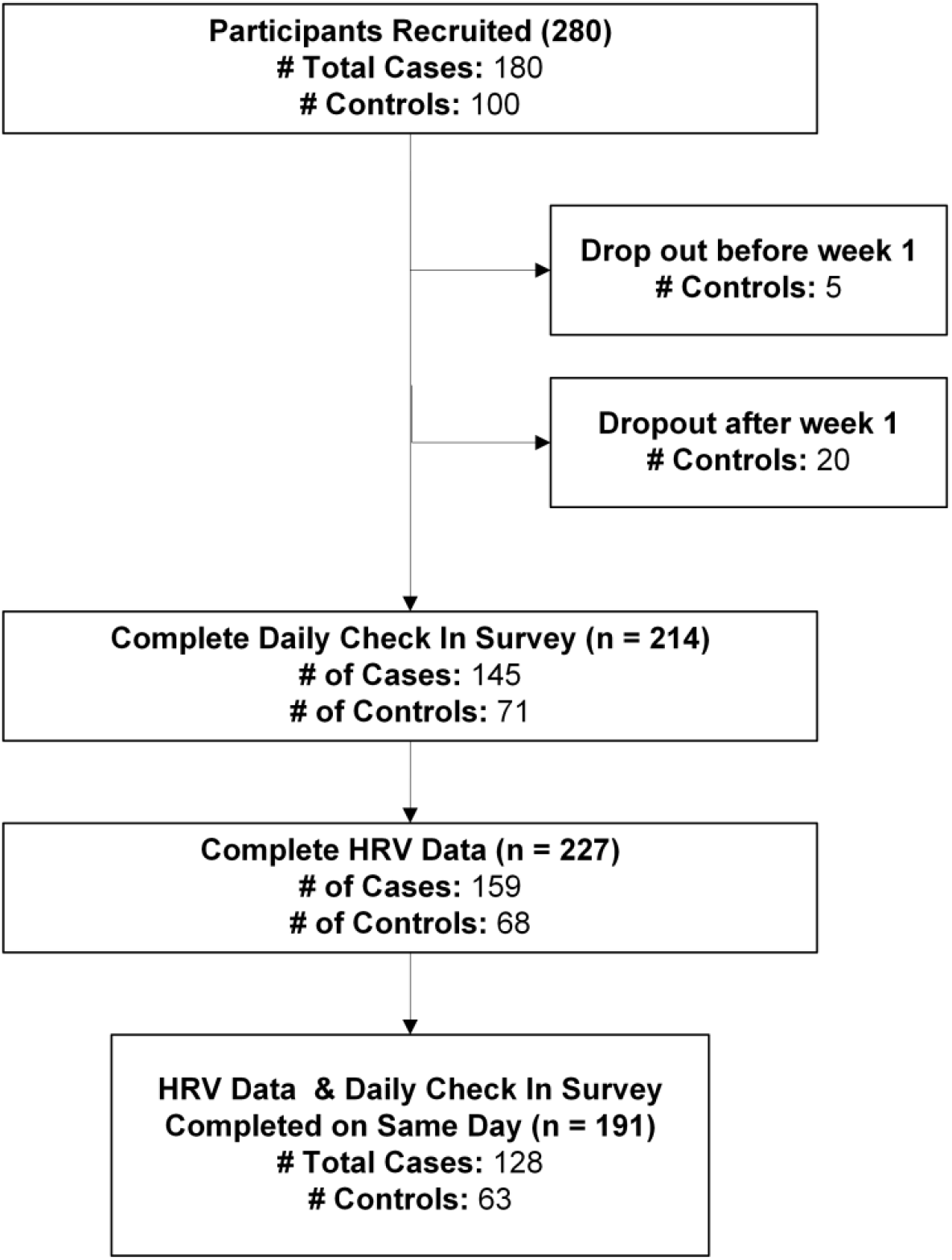
Flow diagram for participant analytical sample

Sleep measure validation / PROMIS Comparison

The following questions were included in the novel daily sleep composite:

1. Was your sleep refreshing?
2. Did you have any problems with your sleep?
3. Did you have difficulty falling asleep?

Each question allowed a response of one of the following, and the associated number value was assigned: none at all (0), a little bit (1), somewhat (2), quite a bit (3), or very much (4). The sleep composite score was found by adding the values of these questions together.

To evaluate convergent validity of the novel daily sleep composite, we regressed PROMIS sleep disturbance T-scores and raw scores on the sleep composite using linear mixed-effects models, using R package lmer ^30^, with random intercepts for participant and study week. Satterthwaite’s method was used for tests. These models accounted for repeated observations within participants and temporal clustering across weeks.

**Supplementary Table 1.**
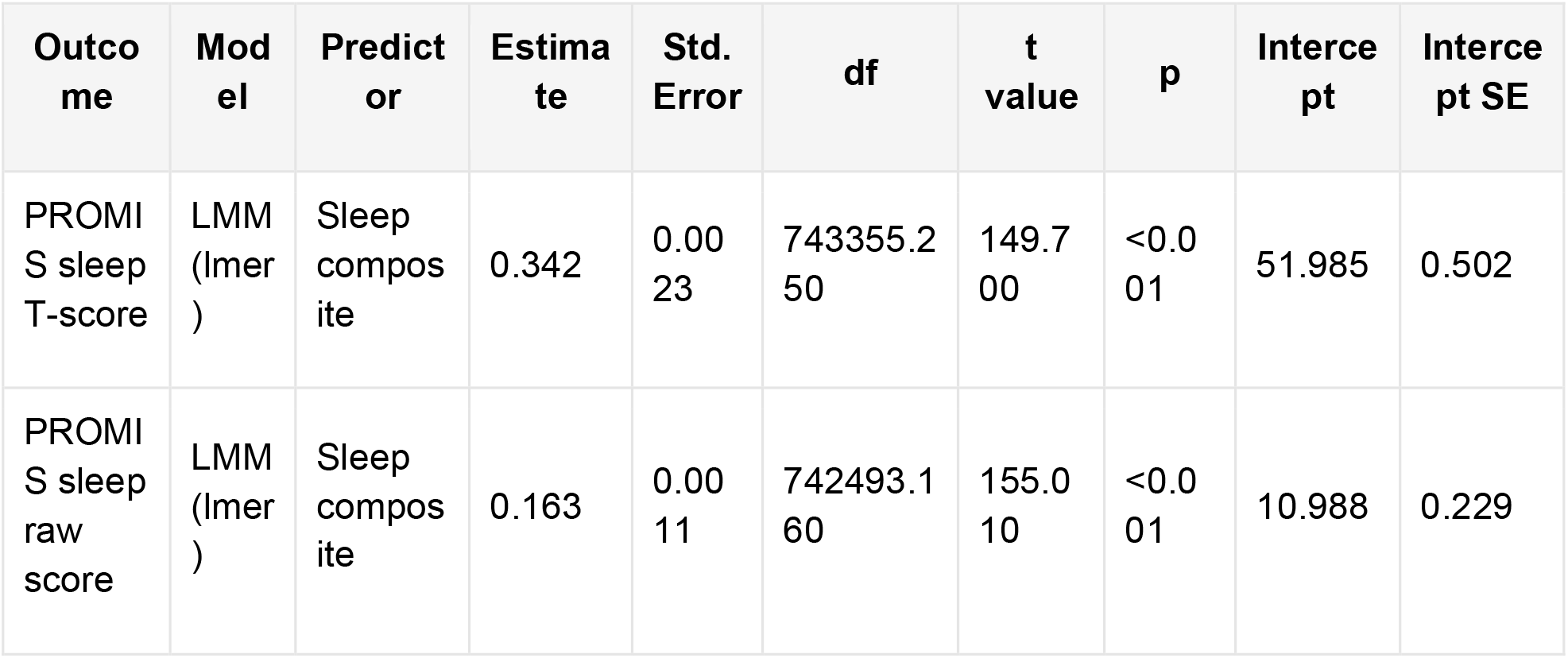
Fixed effects from linear mixed-effects models predicting PROMIS sleep scores from daily composite sleep score.

| Outcome | Model | Predictor | Estimate | Std. Error | df | t value | p | Intercept | Intercept SE |
| --- | --- | --- | --- | --- | --- | --- | --- | --- | --- |
| PROMIS sleep T-score | LMM (lmer) | Sleep composite | 0.342 | 0.0023 | 743355.250 | 149.700 | <0.001 | 51.985 | 0.502 |
| PROMIS sleep raw score | LMM (lmer) | Sleep composite | 0.163 | 0.0011 | 742493.160 | 155.010 | <0.001 | 10.988 | 0.229 |

**Supplementary Table 2.** Parametric terms from generalized additive models predicting PROMIS sleep scores.

| Outcome | Term | Estimate | Std. Error | t value | p value | R<sup>2</sup> (adj.) | Deviance explained (%) |
| --- | --- | --- | --- | --- | --- | --- | --- |
| PROMIS sleep T-score | Sleep Composite | 3.599 | 0.010 | 353.000 | <0.001 | 0.389 | 38.900 |
| PROMIS sleep raw score | Sleep Composite | 1.091 | 0.005 | 233.400 | <0.001 | 0.391 | 39.100 |

In the linear mixed-effects model predicting PROMIS Sleep Disturbance T-score, the novel sleep composite was positively associated with PROMIS score (B = 0.34, SE = 0.002, p < 0.001). The same pattern was observed for PROMIS raw score (B = 0.16, SE = 0.001, p < 0.001). In the GAMM, sleep composite was also positively associated with PROMIS sleep disturbance scores (p<0.001) and had moderate explanatory power, accounting for about 39% variance in PROMIS sleep outcomes. In both models, random intercepts for participant ID and week accounted for repeated measures and temporal clustering. These results indicate that higher values on the novel composite correspond to worse PROMIS sleep disturbance, supporting convergent validity.

**Supplementary Table 3.** Smooth terms from generalized additive models predicting PROMIS sleep scores.

| Outcome | Model | Smooth term | edf | Ref. df | F | p value |
| --- | --- | --- | --- | --- | --- | --- |
| PROMIS sleep T-score | GAM | Sleep Composite | 3.107 | 3.111 | 309079.000 | <0.001 |
| PROMIS sleep T-score | GAM | Week on trial | 0.000 | 1.000 | 0.000 | 0.441 |
| PROMIS sleep T-score | GAM | Participant ID | 1.000 | 1.000 | 4939.000 | <0.001 |
| PROMIS sleep raw score | GAM | Sleep Composite | 3.098 | 3.111 | 28496.000 | <0.001 |
| PROMIS sleep raw score | GAM | Week on trial | 0.754 | 1.000 | 3.137 | 0.042 |
| PROMIS sleep raw score | GAM | Participant ID | 1.000 | 1.000 | 3158.000 | <0.001 |

In supplemental table 3 above, smooth terms indicate that the association between sleep composite and PROMIS sleep score is nonlinear, with effective degrees of freedom are about 3.1 for both outcomes.

## Notes

### Competing Interest Statement

The authors have declared no competing interest.

### Author Declarations

The study protocol was approved by the Institutional Review Board of the Icahn School of Medicine at Mount Sinai, and all participants provided informed consent before enrollment (IRB # STUDY-23-00721).

### Funding

- Eunice Kennedy Shriver National Institute of Child Health & Human Development of the National Institutes of Health: R01HD108263

